# Running acceleration correlates with T2 magnetic resonance imaging values of the lumber intervertebral disc

**DOI:** 10.1101/2024.05.30.24308204

**Authors:** Takayoshi Hakkaku, Yoshiaki Kubo, Koji Koyama, Koichi Nakazato, Takashi Okada, Kenji Hiranuma

## Abstract

**Background:** Running can contribute to both beneficial and detrimental responses in the intervertebral discs (IVDs). To better understand these effects, we investigated the relationship between loading directions during slow running and the rapid changes in T2 times occurring in the lumbar IVDs before and after running.

**Method:** Sixteen healthy male students were fitted with a triaxial accelerator and ran on a treadmill at 8 km/h for one minute. Three lumbar T2 times from the L3/L4 to L5/S1 levels were measured before, immediately after, and 30 minutes post-exercise via magnetic resonance imaging (MRI). The analysis focused on five regions of interest within each disc.

**Result:** Acceleration was 0.23 ± 0.06 root mean square in the mediolateral (X-axis), 1.37 ± 0.08 in the vertical (Y-axis), and 0.30 ± 0.06 in the anteroposterior (Z-axis) direction. A strong correlation was observed between the T2 relaxation times and acceleration, particularly in the Z-axis. At L3/L4, a positive correlation was observed for the posterior nucleus (r = 0.72, p = 0.002, R2 = 0.59). At L4/L5, a positive correlation was observed for the central nucleus (r = 0.73, p = 0.003, R2 = 0.49). At L5/S1, a negative correlation was observed for the anterior annulus fibrosus (r = -0.73, p = 0.01, R2 = 0.48).

**Conclusion:** These results suggest that anteroposterior loading may play a significant role in the response of the IVDs to running.

## Introduction

Human intervertebral discs (IVDs) are primarily avascular tissues. Homeostasis within disc cells depends on nutrient exchange between the disc and the adjacent vertebrae. Nutrients such as glucose and oxygen are transported into the vertebral column via (1) diffusion of nutrients from the vertebral vascular supply terminating at the vertebral endplate and (2) fluid convection due to loading ^1–3^. Oxygen plays a vital role in disc cell metabolism, and the diminished transport of these nutrients is often proposed as a factor in age-related disc degeneration (IDD)^4^.

Quantitative magnetic resonance imaging (MRI) techniques using T2 relaxation times can assess IVDs, and T2 times are strongly correlated with the disc’s biochemical composition ^5^. T2 relaxation times are influenced by two parameters: water content and collagen arrangement in the IVDs. As IDD progresses, the water content decreases, and the collagen arrangement becomes irregular, resulting in decreased T2 relaxation times, particularly within the nucleus pulposus ^6,7^. Investigations using T2 times to elucidate the mechanism of IDD suggest that in the nucleus pulposus, which undergoes fewer degenerative changes, T2 relaxation times are temporarily decreased by exercise stress and recovered by rest ^8^.

Recently, T2 relaxation times have been utilized to examine the beneficial effects of exercise on IVDs. Belavy et al. ^9^ reported that athletic individuals exhibited prolonged T2 relaxation times of lumbar IVDs, whereas long-distance runners exhibited longer T2 values, suggesting greater vertebral heights and IVD hypertrophy. Another study reported longer T2 relaxation times of the IVDs annulus fibrosus and better IDD status in men with a running history of more than 10 years, suggesting that long-term running may delay IDD ^10^. Converesely, Takatalo et al. ^11^ investigated the incidence of IDD in adolescents and reported that running at least twice a week among endurance runners was associated with IDD. This result is inconsistent with earlier studies that proposed that running benefits IVDs. There is no consensus on the effects of running on the intervertebral discs.

Shu et al. conducted a systematic review examining the effects of running on IVDs ^12^. It is widely recognized that rapid deceleration during running could exert significant shock loading on the spine ^13^. Shu et al. reported that the height and volume of IVD reduced after running, attributing this finding to the displacement of fluid in the nucleus pulposus. However, mild to moderate running may lead to super-compensation and improve IVD parameters in habitual runners. Additionally, 0.2-0.8 MPa has been reported as the optimal load range to induce an anabolic response in IVDs ^14^. In support of this, Wilke et al. posit that activities ranging from walking to slow running and/or quickly walking fall within that optimal range ^15^. Belavy et al. ^9^ used a triaxial accelerometer and measured the mean amplitude deviation (MAD) of acceleration during running. They found that 0.44 to 0.59 g is optimal for lumbar IVDs. In addition, the previous study showed that ambulation at 7.2 km/h fell within this 0.44 and 0.59 *g* MAD range. In other words, activities ranging from quickly walking (more than 5.4 km/h) to slow running (less than 9 km/h) may benefit the discs. However, activities outside of this range may not yield positive effects.

However, Min et al. reported a lower prevalence of disc IDD in track and field athletes than in the general population ^16^. Given that track and field athletes run at speeds outside the optimal loading range to induce an anabolic response in IVDs, we believe that speed alone cannot fully account for the observed effect on IVDs and that factors other than speed need to be considered. Although Belavy et al. employed MAD to assess the load on IVDs during running, which is calculated from acceleration in three directions (anteroposterior, medial-lateral, and vertical), it remains unclear which specific direction exerts the most influence on the IVD fluid content during running.

Therefore, this study aimed to determine which direction of acceleration exerts the most significant effect on IVD fluid content by examining acceleration during running and the displacement of IVD fluid before and after running. We hypothesize that by investigating the acceleration profiles during running at speeds considered to be within the optimal range for IVDs, valuable insights for improving T2 times and preventing IDD could be obtained. Our primary hypothesis posits that anteroposterior acceleration may account for the changes observed in IVD fluid content before and after running. Given that running primarily occurs in the sagittal plane, and Wilke et al. reported significant changes in IVD internal pressure in flexion-extension ^15^, the influence of anteroposterior loading on IVD requires further investigation. While an increase in running speed requires an increase in vertical ground force ^17^, vertical acceleration is unlikely to be the most relevant factor for IVD fluid content. This assertion is supported by the low incidence of IDD in track and field athletes ^16^.

## Materials and methods

### Participants

The local ethics committee approved this study (no. 022-H173; January 31, 2023), which adhered to the guidelines for experimental studies involving human participants and met the ethical standards of the journal. The study was conducted following the principles of the Declaration of Helsinki. All participants were informed of the study’s purpose, experimental procedures, potential benefits, and possible risks and were enrolled after providing written informed consent.

As noted in the introduction, there is no consensus on the effect of running on IVDs. Therefore, to establish a baseline for comparison, we first identified the characteristics of non-runners. Additionally, sixteen healthy male university students were enrolled in this study (age = 20.8 ± 1.2 years; height = 172.2 ± 4.4 cm; weight = 72.3 ± 9.2 kg; mean ± standard deviation). The participants had no history of low back pain and did not specialize in long-distance running. Low back pain was assessed using a 10-question questionnaire (OCU-test) on LBP related to activities of daily living, developed by Osaka City University and modified by Kuroki and Tajima ^18^. All participants were instructed to respond based on their status, and all were judged to be without low back pain. The severity of IDD was also assessed from T2-weighted images in the sagittal plane; grade 3 or higher was defined as IDD according to the classification of Pfirrmann et al. ^19^, and applicable IVDs were excluded. Orthopaedic surgeons specializing in spine disorders, blinded to the participants’ injury status, assessed the MRI scans.

### Procedures

Upon arrival at the laboratory, participants received a comprehensive explanation of the study, completed a baseline questionnaire (duration: 10 minutes), were placed on their backs in bed and allowed to rest for 10 minutes before pre-MRI testing (Pre). Thereafter, patients were fitted with a triaxial accelerometer and performed a 1-minute run, followed by post-MRI testing (Post). After running, the participants remained in a supine position throughout the post-exercise period, and MRI measurements were obtained 30 minutes after running to determine IVD changes over time (Post 30).

### Running exercise and triaxial accelerometer

The participants engaged in running trials on a treadmill (Elite 5000; Johnson, Tokyo, Japan). The running speed was set at 8 km/h, within the MAD range reported by Belavy et al., which positively impacted the IVDs ^9^. The speed was gradually increased to the target velocity of 8 km/h, and the participants ran for 1 minute at this speed. The exercise duration in this study was informed by the findings of Chokan et al., who reported that T2 values changed even with minimal exercise loads and duration (15 repetitions of trunk flexion, extension, and rotation exercises) ^8^. The participants were instructed to maintain their usual running style while wearing their preferred running shoes. No warm-up was performed before the run, and the experiment began as soon as the participants were ready.

To measure the load caused by running, a triaxial accelerometer (wGT3X-BT; ActiGraph, Pensacola, FL, USA) with a range of ± 8.0 g was employed. A triaxial accelerometer was fixed to the L4-L5 lumbar intervertebral region using a belt by medically qualified staff. The x-axis was oriented in the medial-lateral direction, the y-axis in the vertical direction, and the z-axis in the anteroposterior direction. Positive X, Y, and Z values represented the right, downward and backward acceleration, respectively. Raw acceleration data were collected at 100 Hz during the treadmill running task, transferred to analysis software (OT Biolab; OT Bioelettronica, Torino, Italy) and filtered at a cut-off frequency of 10 Hz ^20^ . The RMS 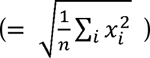 of the x-axis (ML-RMS), y-axis (VT-RMS), and z-axis (AP-RMS), resultant acceleration ( = 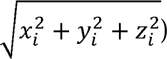 and MAD ( = 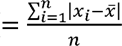, *x* = resultant acceleration) were calculated at 30 s, excluding the 15 s before and after the 1-minute running trials, respectively.

### Testing and MRI scanning protocol

The participants were instructed not to perform any exercise on the day of the scan. T2 times before and after exercise were measured using MRI (ECHELON OVAL, 1.5 T; Hitachi Medical Systems, Tokyo, Japan). To mitigate the effects of normal diurnal variations on the spine ^21^, all tests were performed at approximately noon. The resulting images were evaluated by an orthopedic surgeon specializing in spinal diseases to assess the degree of IDD.

Sagittal T2 mapping using spin-echo multi-echo sequences (nine echo times: 12, 24, 36, 48, 60, 72, 84, 96, and 108 ms; repetition time, 1600 ms; number of slices, 12; slice thickness, 4 mm; gap, 5 mm; interslice distance, 5.0 mm; field of view, 200 mm; resolution, 1.04 × 1.25 × 4.00 mm per pixel; and acquisition time, 6 minutes 55 s). □

T2 times were calculated using the MRI’s built-in analysis program. After the images were obtained in ECHELON OVAL format, they were loaded and viewed using the T2*RelaxMap feature of the MRI system (Hitachi Medical Systems). After the images of the nine echo times were superimposed using the software, the outlines of the lumbar IVDs were manually traced to determine the region of interest (ROI) in each image. T2 times for each pixel were calculated using the software, whereas T2 times were measured for each slice. The three IVDs analyzed were L3/L4 to L5/S1, which had a high incidence of IDD. The IVDs were manually segmented into five subregions from anterior to posterior aspect (Fig 1) ^22^: from front to back, the anterior annulus fibrosus (AF), anterior nucleus (AN), central nucleus (CN), posterior nucleus (PN), and posterior annulus fibrosus (PF). Image calculations were performed thrice for each ROI, and the average values were used as T2 times. The intraclass correlation coefficient values ranged from 0.84 to 1.00, suggesting that the measurement methods used in this study are highly reproducible (Supplementary material Table 1).

**Fig 1.**
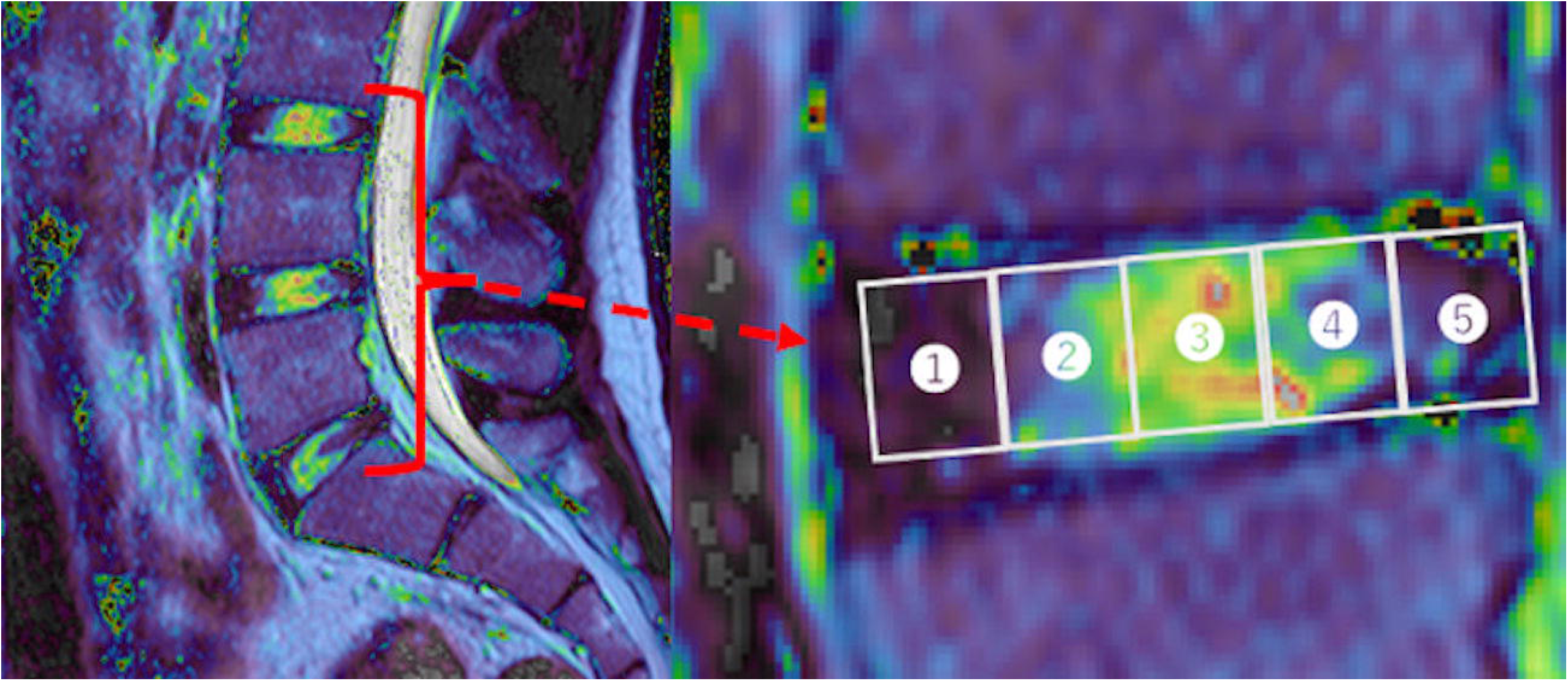
Examples of IVDs and ROIs on MR images. T2* RelaxMap of lumbar intervertebral discs in the central slice (left) and from L3/L4 to L5/S1 divided into five subregions (right): 1. anterior annulus fibrosus; 2. anterior nucleus; 3. Central nucleus; 4. posterior nucleus; and 5. posterior annulus fibrosus.

### Statistical analysis

Data are expressed as meanL±Lstandard deviation. T2 relaxation times for each level and each site at Post and Post 30 were normalized to the corresponding T2 values for the same levels and sites at Pre. The normalized T2 times were used for the analysis.

One-way analysis of variance (ANOVA) was used to examine the changes in the normalized T2 relaxation times over time at each level and site.

Pearson’s correlation coefficient (*r*) was used to examine the correlation between the normalized T2 times for each level and each site at each time point, the root mean square values (RMS) for acceleration in the mediolateral (ML-RMS), vertical (VT-RMS), anteroposterior (AP-RMS), resultant acceleration, and MAD. The central nucleus (CN) at Post 30 in L3/4, the posterior annulus fibrosus (PF) at Post in L4/5, the PF at Post in L5/S1 and the anterior nucleus (AN) at Post 30 were not normally distributed. Therefore, they were calculated using the nonparametric Spearman correlation coefficient (ρ). In addition, we also estimated the statistical power of individual outcomes by using the G∗Power software version 3.1 with an alpha level of 5% to determine to what extent a statistical difference between the two groups of results can be detected ^23^. Where a correlation was found and statistical power was 0.8 or greater, a simple linear regression analysis was further performed to determine adjusted R^2^. All tests were performed using the statistical analysis software SPSS Statistics for Macintosh (ver. 29.0.2.0; IBM, Armonk, NY, USA). The significance level was set at p < .05.

## Results

### T2 times

Of the 16 participants in the study, two discs were classified as Pfirrmann grade III on L4/L5, and four discs were Pfirrmann grade III on L5/S1. Therefore, 3 of the 48 IVDs were excluded, leaving 45 IVDs for inclusion. The T2 times for each IVD level are shown in Table 1. One-way repeated measures analysis of variance revealed no significant differences in the changes over time in any of the ROI. The normalized T2 times for each IVD are shown in Table 2.

**Table 1.** T2 relaxation times within the intervertebral disc pre-versus post-running.

|  |  | pre(ms) | post(ms) | post30(ms) |
| --- | --- | --- | --- | --- |
|  | AF | 101.7 ± 26.6 | 101.8 ± 25.3 | 100.2 ± 25.6 |
|  | AN | 165.5 ± 34.1 | 165.2 ± 33.5 | 163.4 ± 38.1 |
| L3/L4 | CN | 210.9 ± 25.8 | 209.3 ± 19.4 | 209.7 ± 24.5 |
|  | PN | 165.6 ± 32.8 | 158.2 ± 23.2 | 160.8 ± 24.2 |
|  | PF | 94.3 ± 29.2 | 90.4 ± 18.9 | 89.0 ± 18.0 |
|  | AF | 97.6 ± 28.4 | 96.7 ± 27.2 | 96.6 ± 23.3 |
|  | AN | 165.2 ± 44.5 | 158.0 ± 38.4 | 158.6 ± 31.0 |
| L4/L5 | CN | 224.2 ± 30.4 | 219.3 ± 34.5 | 219.8 ± 37.5 |
|  | PN | 173.3 ± 31.3 | 171.5 ± 29.0 | 175.2 ± 35.0 |
|  | PF | 97.6 ± 24.0 | 104.2 ± 31.7 | 95.2 ± 25.1 |
|  | AF | 101.0 ± 30.7 | 105.6 ± 28.7 | 101.6 ± 22.8 |
|  | AN | 154.1 ± 46.2 | 155.7 ± 44.3 | 149.8 ± 32.1 |
| L5/S1 | CN | 193.5 ± 55.0 | 191.2 ± 54.4 | 193.4 ± 45.1 |
|  | PN | 143.7 ± 36.2 | 141.0 ± 29.0 | 148.5 ± 35.2 |
|  | PF | 88.3 ± 17.8 | 86.1 ± 26.6 | 92.4 ± 18.2 |

**Note:** Data are shown as mean ± standard deviation.
**Abbreviations:** AF, annulus fibrosus; AN, anterior nucleus; CN, central nucleus; PN, posterior
nucleus; PF, posterior annulus fibrosus; Post, post-running; Post30, 30 min post-running; Pre,
pre-running; ROI, region of interest

**Table 2.**
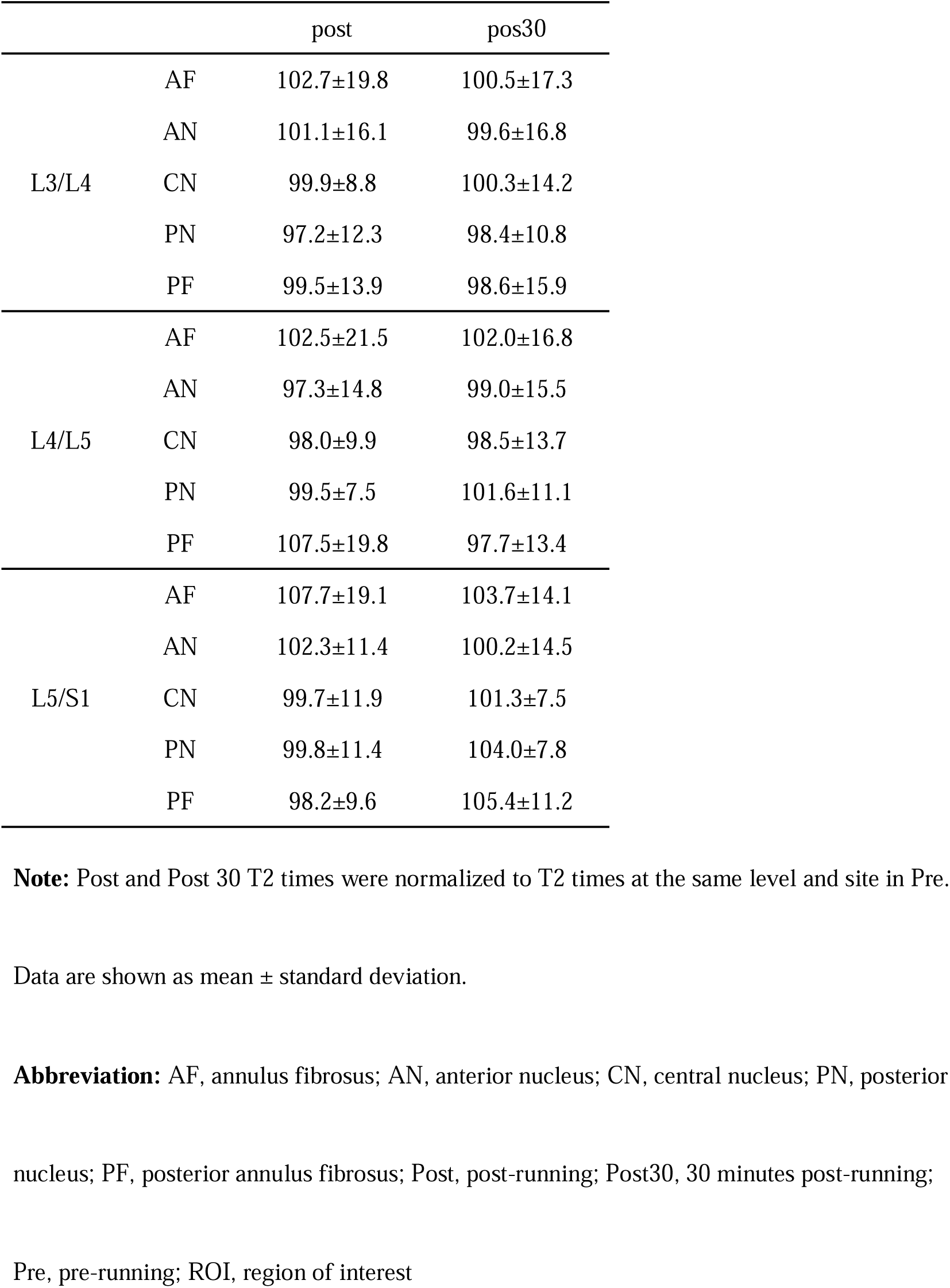
The normalized T2 times.

|  |  | post | pos30 |
| --- | --- | --- | --- |
| L3/L4 | AF | 102.7±19.8 | 100.5±17.3 |
|  | AN | 101.1±16.1 | 99.6±16.8 |
|  | CN | 99.9±8.8 | 100.3±14.2 |
|  | PN | 97.2±12.3 | 98.4±10.8 |
|  | PF | 99.5±13.9 | 98.6±15.9 |
| L4/L5 | AF | 102.5±21.5 | 102.0±16.8 |
|  | AN | 97.3±14.8 | 99.0±15.5 |
|  | CN | 98.0±9.9 | 98.5±13.7 |
|  | PN | 99.5±7.5 | 101.6±11.1 |
|  | PF | 107.5±19.8 | 97.7±13.4 |
| L5/S1 | AF | 107.7±19.1 | 103.7±14.1 |
|  | AN | 102.3±11.4 | 100.2±14.5 |
|  | CN | 99.7±11.9 | 101.3±7.5 |
|  | PN | 99.8±11.4 | 104.0±7.8 |
|  | PF | 98.2±9.6 | 105.4±11.2 |

**Note:** Post and Post 30 T2 times were normalized to T2 times at the same level and site in Pre.
Data are shown as mean ± standard deviation.
**Abbreviation:** AF, annulus fibrosus; AN, anterior nucleus; CN, central nucleus; PN, posterior
nucleus; PF, posterior annulus fibrosus; Post, post-running; Post30, 30 minutes post-running;
Pre, pre-running; ROI, region of interest

### Triaxial acceleration during running

The ML-RMS was 0.23 ± 0.06 g, VT-RMS was 1.37 ± 0.08 g, and AP-RMS was 0.30 ± 0.06 g, respectively. The resultant acceleration and MAD were 1.43 ± 0.07 g and 0.71 ± 0.09 g, respectively.

### Correlation between the normalized T2 times and triaxial acceleration

The results of the correlation analysis between the normalized T2 times and acceleration are shown in Tables 3 and 4. In terms of the relationship between the normalized T2 times at Post and triaxial acceleration, ML-RMS was related to the normalized T2 times of AN (r = -0.58, p = 0.02, statistical power = 0.69) and CN (r = -0.54, p = 0.03, statistical power = 0.61) in L3/L4. VT-RMS was related to the normalized T2 times of PF (r = -0.57, p = 0.02, statistical power = 0.67) in L3/L4. AP-RMS was related to the normalized T2 times of AN (r = 0.53, p = 0.04, statistical power = 0.59) and CN (r = 0.62, p = 0.01, statistical power = 0.77) in L3/L4, and the normalized T2 times of CN (r = 0.73, p = 0.003, statistical power = 0.89), and PF (r = -0.59, p = 0.03, statistical power = 0.64) in L4/L5, and the normalized T2 times of AF (r = -0.73, p = 0.01, statistical power = 0.83) and PN (r = -0.59, p = 0.03, statistical power = 0.56) in L5/S1. Resultant acceleration was related to the normalized T2 times of PF (r = 0.55, p = 0.03, statistical power = 0.63) in L3/L4. MAD was related to the normalized T2 times of PF (r = 0.68, p = 0.004, statistical power = 0.87) in L3/L4.

**Table 3.**
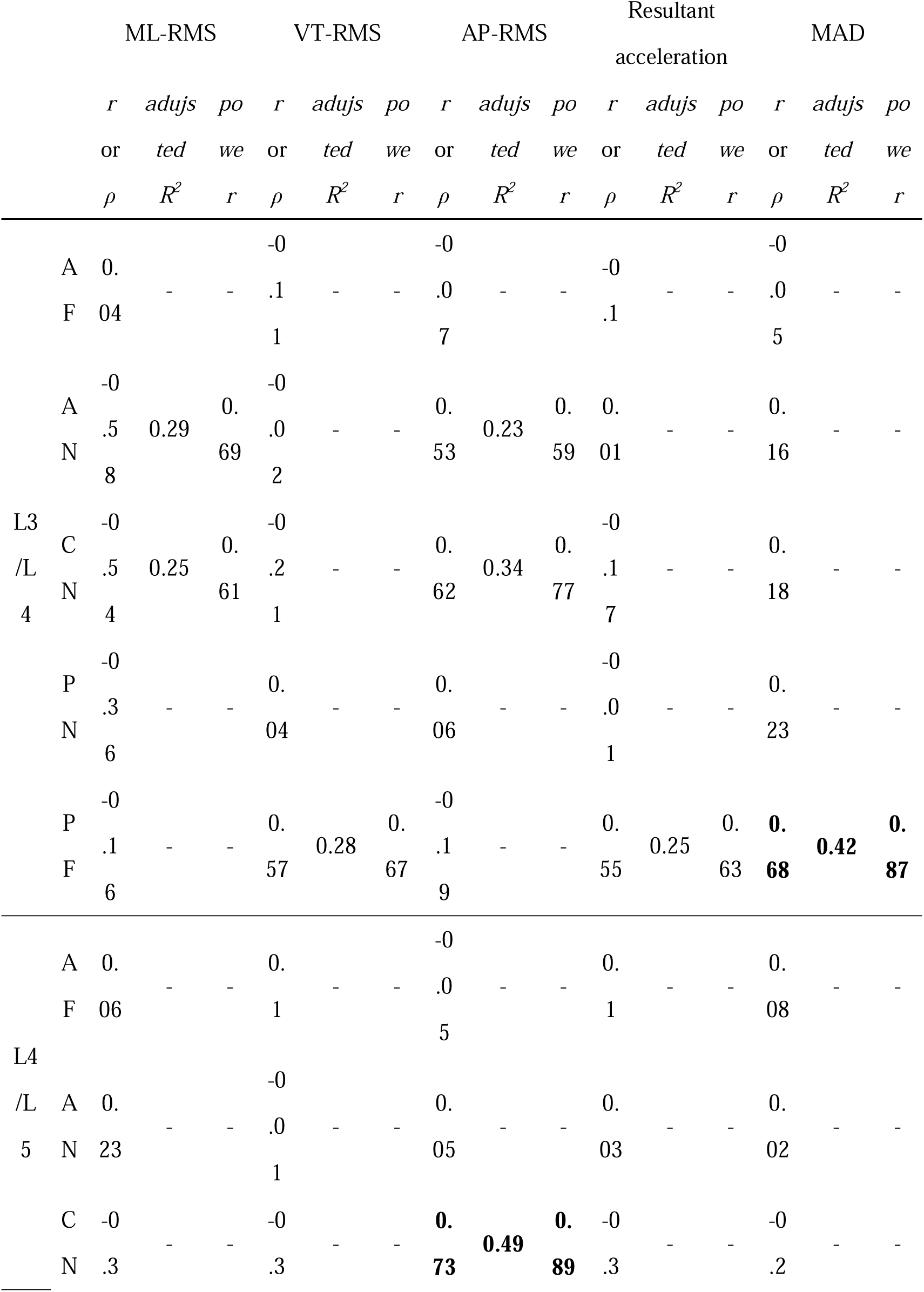
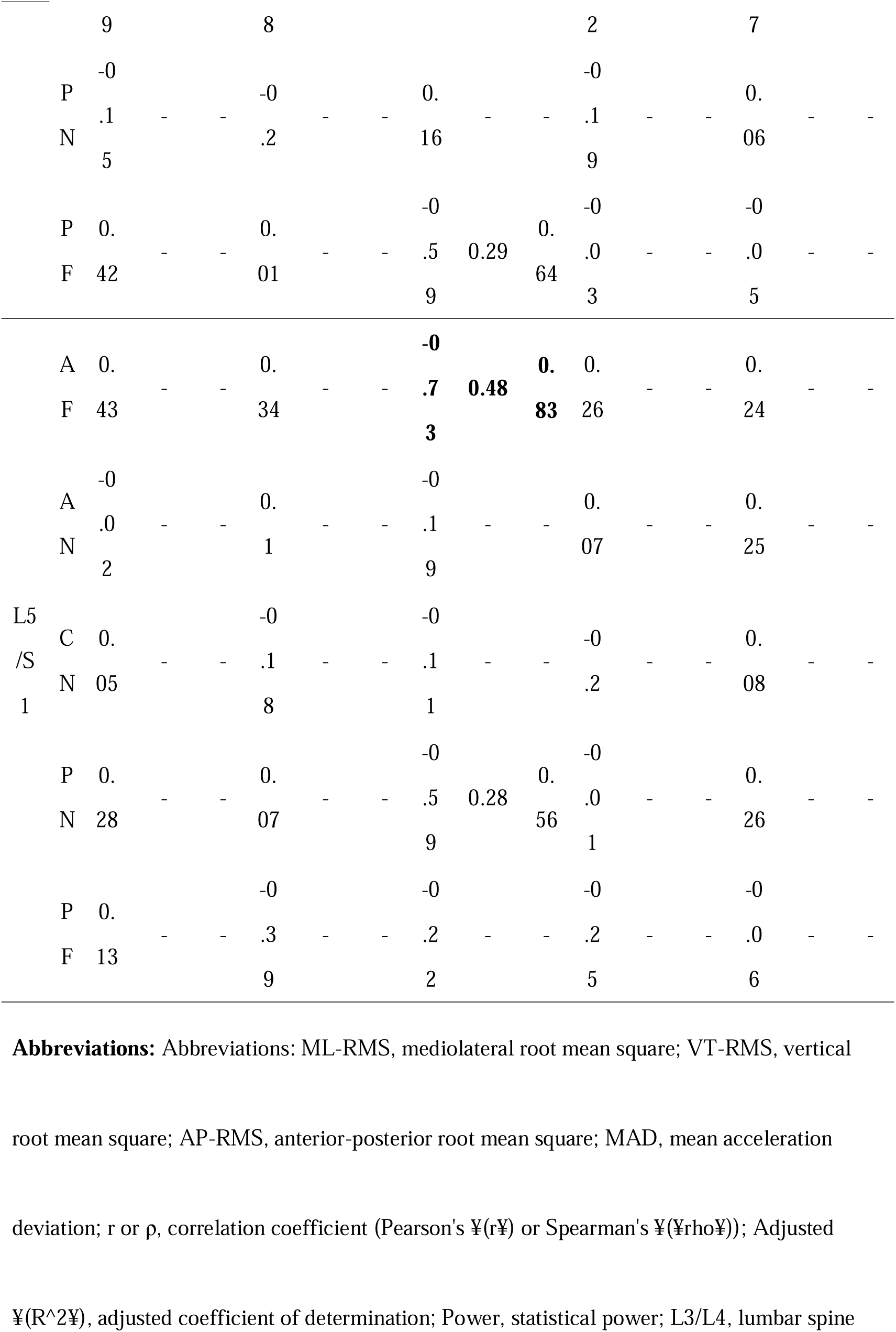
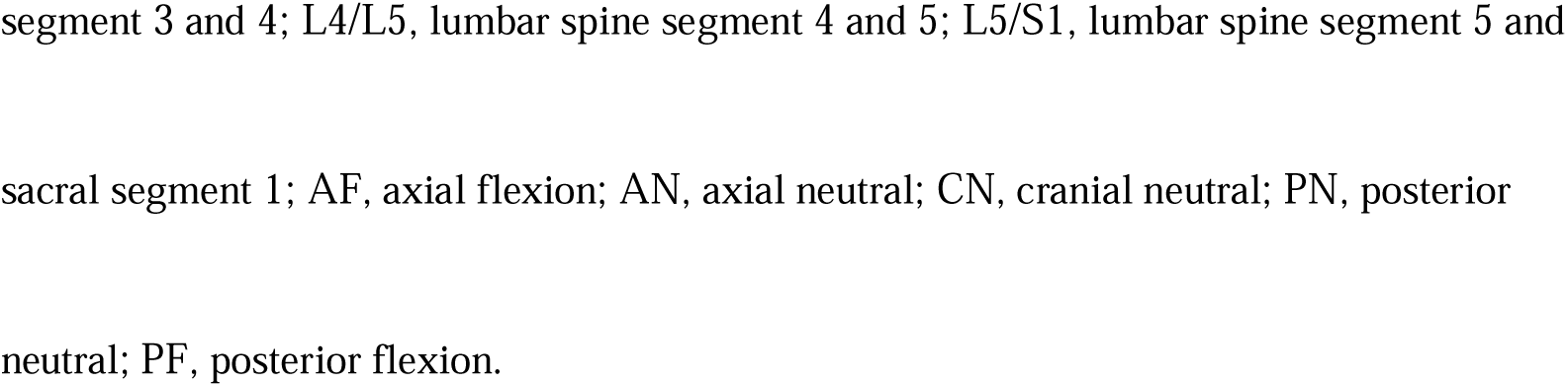
Correlation between post-T2 relaxation times and triaxial acceleration.

**Table 4.**
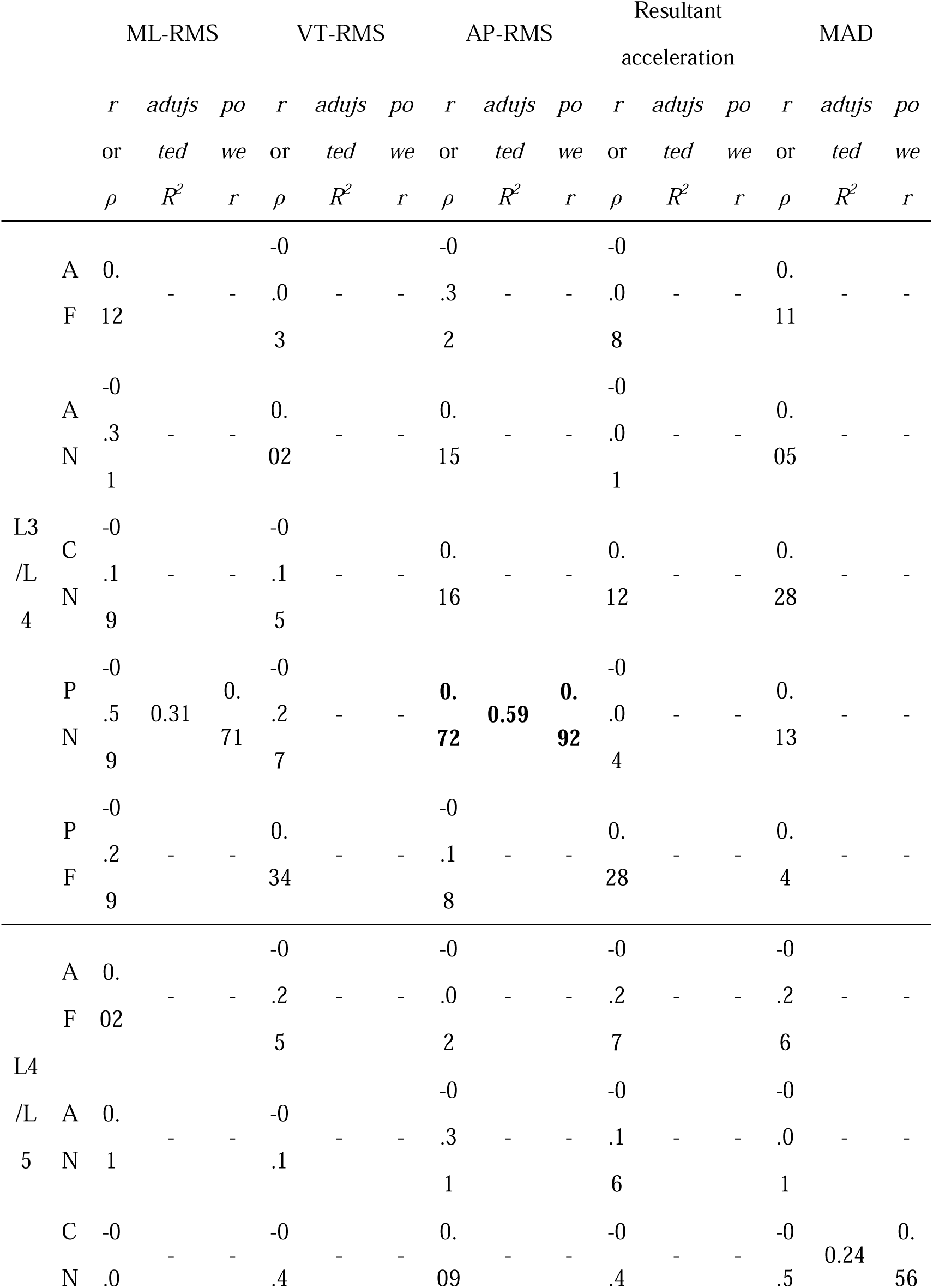
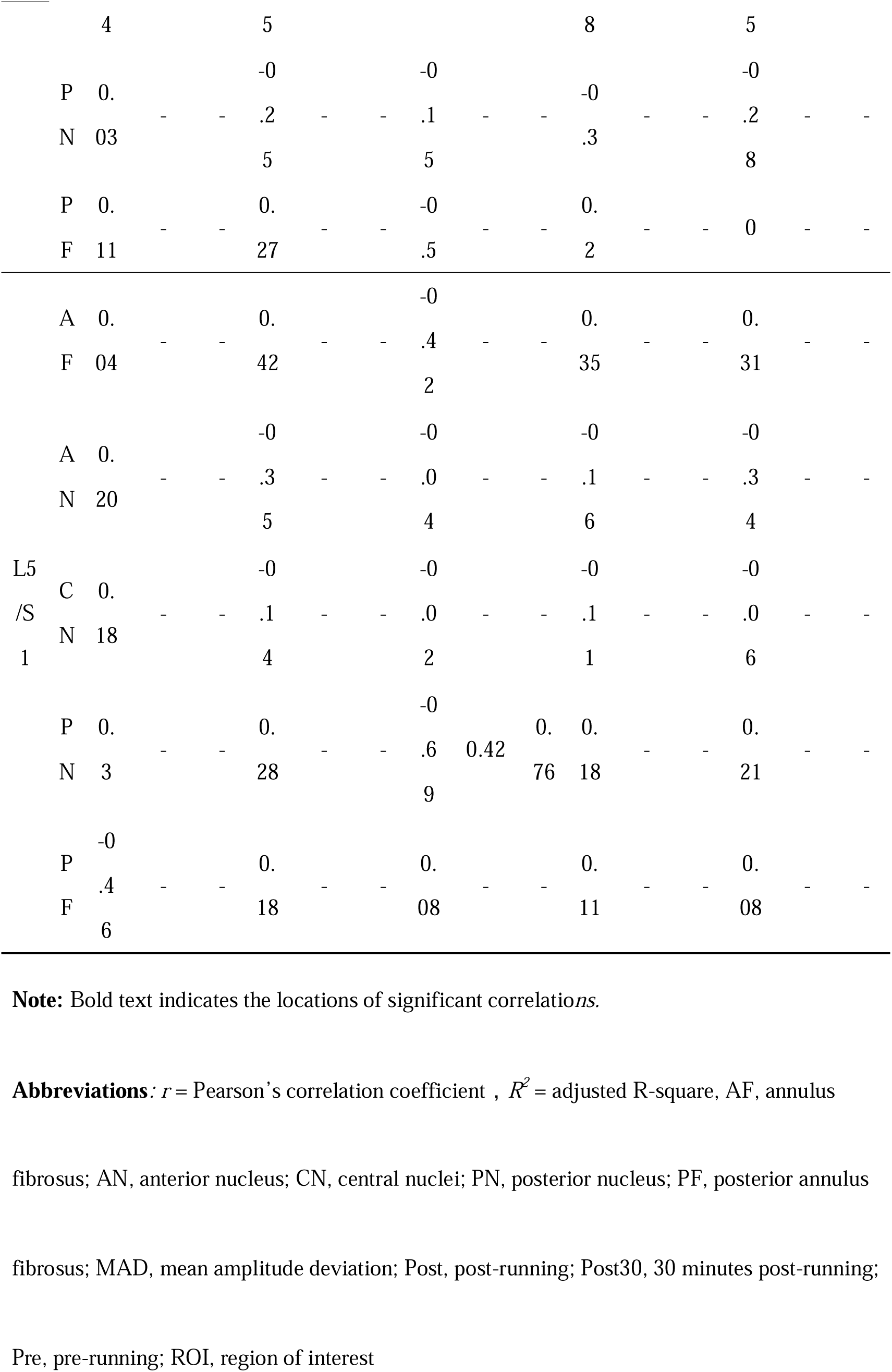
Correlation between post30 T2 relaxation times and triaxial acceleration.

In terms of the relationship between the normalized T2 times at Post 30 and triaxial acceleration, ML-RMS was related to the normalized T2 times of PN (r = -0.59, p = 0.02, statistical power = 0.71) in L3/L4. AP-RMS was related to the normalized T2 times of PN (r = 0.72, p = 0.002, statistical power = 0.92) in L3/L4, and the normalized T2 times of PN (r = -0.69, p = 0.01, statistical power = 0.76) in L5/S1. MAD was related to the normalized T2 times of CN (r = -0.55, p = 0.04, statistical power = 0.56) in L4/L5.

Since correlations were observed between the normalized T2 times of CN at Post in L4/L5 and AP-RMS, and between the normalized T2 times of AF at Post in L5/S1 and the AP-RMS, between the normalized T2 times of PF at Post in L3/L4 and MAD, and between the normalized T2 times of PN at Post 30 in L3/L4 and the AP-RMS and there was sufficient statistical power, a simple linear regression analysis was performed to investigate the associations. The regression models were as follows: the normalized T2 times of CN at Post in L4/L5 = 115.4 + 63.2 × AP-RMS (adjusted R^2^ = 0.49), the normalized T2 times of AF at Post in L5/S1 = 219.9 + 174.1 × AP-RMS (adjusted R^2^ = 0.48), the normalized T2 times of PF at Post in L3/L4 = 104.6 + 25.6 × MAD (adjusted R^2^ = 0.42), and the normalized T2 times of PN at Post 30 in L3/L4 = 144.5 + 55.2 × AP-RMS (adjusted R^2^ = 0.59).

## Discussion

There is no unified consensus on the effects of running load on IVDs. This study investigated the relationship between the change in T2 times before and after slow running, which is considered to be suitable for IVDs, and the direction of loading during running. The results revealed that in slow running in young, healthy individuals without a prior history of regular running, the correlation between the change in T2 times and the direction of loading was different for each IVD level. In particular, the findings suggest that acceleration of anteroposterior direction exerts the most significant influence on T2 times after running.

In this study, an optimal speed range of 8 km/h and the running time were determined based on the reports of Chokan et al. and Bellaby et al. ^8,9^. Chokan et al. showed that in the nucleus pulposus, T2 times significantly decreased after exercise and returned to the pre-exercise level after a recovery period. The exercise protocol selected by Chokan et al. involved performing 15 repetitions of trunk movements, including 30° extension, 45° flexion, and 40° left and right rotation. The exercise time lasted between 1-2 minutes. Therefore, it was hypothesized that T2 times within the nucleus pulposus would exhibit significant changes pre- and post-running in this study. However, there were no changes at any region of interest (ROI) or IVD level before and after the 1-minute run at 8 km/h (Table 1). It is speculated that the range of motion (ROM) during running in this study was narrower than the exercises employed in the previous study by Chokan et al. Thus, this suggests that the direction of motion and ROM of the torso may be related to T2 times.

Belavy et al. reported that running exerts the most significant impact on T2 times within the nucleus pulposus. In addition, Belavy et al. mentioned that MAD in the range of 0.44 to 0.59 g during running at speeds of 5.4 to 9 km/h has a positive effect on IVDs ^8,9^. Regarding the relationship between nucleus pulposus and acceleration, MAD is negatively correlated with the normalized T2 time of CN at Post 30 in L4/L5 (r = -0.55, statistical power = 0.56). Although the running speed employed in this study was 8 km/h, the MAD was 0.71 ± 0.09 g, exceeding the range required for a positive effect on the IVDs. This result was consistent with previous study ^9^. However, MAD alone cannot explain the normalized T2 times of the nucleus pulposus after running due to the limited statistical power in this analysis. No correlation was observed between the acceleration in the vertical direction and IVD of

CN at each level. In contrast, the direction of motion might impact the T2 times. A positive correlation was revealed between AP-RMS and the normalized T2 times of CN at Post in L3/L4 (r = 0.62, statistical power = 0.77) and L4/L5 (r = 0.73, statistical power = 0.89). Thus, it was found that higher anteroposterior acceleration during running is associated with higher IVD fluid content immediately after running. In particular, the change in the fluid content of the nucleus pulposus in L4/L5 immediately after running related to approximately 50% of the anteroposterior acceleration (adjusted R^2^ = 0.49). During trunk motion, the greatest impact on intradiscal pressure is in flexion-extension ^24^, as it produces a range of 0.3-1.2 MPa, the optimal pressure for IVDs during the movement. Meadows et al. also suggest that flexion is a more important loading condition for mechanical function (axial strain, disc wedge angle changes, and anterior-posterior shear displacement) of the disc in vivo, according to a mechanical study of IVDs ^25^. These factors support the hypothesis that anteroposterior loading significantly affects the fluid content of the nucleus pulposus in L3/L4, and L4/L5.

In contrast, for the IVD of L5/S1, no correlation was observed between the nucleus pulposus and the anteroposterior acceleration. However, a negative correlation was found between the annulus fibrosus and the anteroposterior acceleration. The IVD of L5/S1 had greater posterior shear forces due to flexion than the other levels ^25^. Shear forces may induce IDD ^26,27^. Therefore, regarding the IVD of L5/S1, the flexion-extension force becomes a shear force, which may have reduced the fluid content of the annulus fibrosus.

Interestingly, a positive correlation was observed between normalized T2 times of PF in L3/L4 and MAD (r = 0.68, statistical power = 0.87, adjusted R^2^ = 0.42). Although the statistical power is limited, these findings suggest that the IVD of L3/L4 may be affected by acceleration not only in the anterior-posterior direction but also in the lateral and vertical directions. A high prevalence of upper IDD has been reported in gymnasts with high rotational stress ^28,29^. Given that the IVD in L3/L4 exhibits greater lateral flexion and rotation than the lower levels, and is loaded in various directions, it is hypothesized that these multi-directional forces are indicated in the MAD values, representing the deviation of acceleration in each axis.

Based on the present results, the hypothesis that slow running improves T2 times of the nucleus pulposus was rejected because it did not significantly change T2 times. However, the results partially support the hypothesis because it improved anteroposterior loading on T2 values. This study had several limitations. First, a treadmill was used to maintain a constant running speed within the optimal loading range of the IVDs. However, the similarity in the triaxial acceleration data between treadmill and ground running suggests that the findings for the loading direction were unaffected ^30^. Second, although only male participants were included in the study, sex was standardized because sex differences have been observed in pelvic movements during running ^31^, which may influence loading patterns. Finally, although a correlation was observed, other factors might contribute to IDD. Further research is needed to investigate factors other than speed that may contribute to IDD. In addition, this study focused on immediate post-exercise changes, and whether its findings can be applied to IDD prevention strategies is an issue to be examined. Further investigations should incorporate slow-running motion analysis to determine the reasons for the observed high anteroposterior loading. Despite these limitations, the evidence suggests that IVDs exhibit a beneficial response to certain types of loading and may have public health implications. From the results of the present study, we believe that other exercise interventions could also benefit from the optimization of loading parameters. Given the critical role of IVDs in spinal pain development, understanding how IVDs respond to certain types of stress will lead to improved exercise guidelines for the prevention and management of spinal pain.

## Conclusion

The results of this study indicate that changes in T2 times of the IVDs while running at 8 km/h correlate with acceleration, suggesting that anteroposterior acceleration may contribute to the important adaptive response within the IVDs.

## Data Availability

All relevant data are within the manuscript and its Supporting Information files.

## Data Availability Statement

The data that support the findings of this study are available on request from the corresponding author. The data are not publicly available due to privacy or ethical restrictions.

## Acknowledgements

The authors would like to express their sincere gratitude to all members of the research team and all participants who contributed to this study.

## References

1. Huang YC, Urban JP, Luk KD. Intervertebral disc regeneration: do nutrients lead the way? Nat Rev Rheumatol. Sep 2014;10(9):561–6. doi:10.1038/nrrheum.2014.91

2. Urban JP, Holm S, Maroudas A, Nachemson A. Nutrition of the intervertebral disk. An in vivo study of solute transport. Clin Orthop Relat Res. Nov-Dec 1977;(129):101–14.

3. Urban JP, Smith S, Fairbank JC. Nutrition of the intervertebral disc. Spine (Phila Pa 1976). Dec 1 2004;29(23):2700–9. doi:10.1097/01.brs.0000146499.97948.52

4. Grunhagen T, Wilde G, Soukane DM, Shirazi-Adl SA, Urban JP. Nutrient supply and intervertebral disc metabolism. J Bone Joint Surg Am. Apr 2006;88 Suppl 2:30–5. doi:10.2106/JBJS.E.01290

5. Huang L, Liu Y, Ding Y, et al. Quantitative evaluation of lumbar intervertebral disc degeneration by axial T2∗ mapping. Medicine. 2017;96(51)

6. Marinelli NL, Haughton VM, Anderson PA. T2 relaxation times correlated with stage of lumbar intervertebral disk degeneration and patient age. AJNR Am J Neuroradiol. Aug 2010;31(7):1278–82. doi:10.3174/ajnr.A2080

7. Stelzeneder D, Welsch GH, Kovacs BK, et al. Quantitative T2 evaluation at 3.0T compared to morphological grading of the lumbar intervertebral disc: a standardized evaluation approach in patients with low back pain. Eur J Radiol. Feb 2012;81(2):324–30. doi:10.1016/j.ejrad.2010.12.093

8. Chokan K, Murakami H, Endo H, et al. Evaluation of Water Retention in Lumbar Intervertebral Disks Before and After Exercise Stress With T2 Mapping. Spine (Phila Pa 1976). Apr 2016;41(7):E430–6. doi:10.1097/BRS.0000000000001283

9. Belavy DL, Quittner MJ, Ridgers N, Ling Y, Connell D, Rantalainen T. Running exercise strengthens the intervertebral disc. Sci Rep. Apr 19 2017;7:45975. doi:10.1038/srep45975

10. Mitchell UH, Bowden JA, Larson RE, Belavy DL, Owen PJ. Long-term running in middle-aged men and intervertebral disc health, a cross-sectional pilot study. PLoS One. 2020;15(2):e0229457. doi:10.1371/journal.pone.0229457

11. Takatalo J, Karppinen J, Nayha S, et al. Association between adolescent sport activities and lumbar disk degeneration among young adults. Scand J Med Sci Sports. Dec 2017;27(12):1993–2001. doi:10.1111/sms.12840

12. Shu D, Dai S, Wang J, Meng F, Zhang C, Zhao Z. Impact of Running Exercise on Intervertebral Disc: A Systematic Review. Sports Health. Nov-Dec 2024;16(6):958–970. doi:10.1177/19417381231221125

13. Giandolini M, Horvais N, Rossi J, Millet GY, Samozino P, Morin JB. Foot strike pattern differently affects the axial and transverse components of shock acceleration and attenuation in downhill trail running. J Biomech. Jun 14 2016;49(9):1765–1771. doi:10.1016/j.jbiomech.2016.04.001

14. Belavy DL, Albracht K, Bruggemann GP, Vergroesen PP, van Dieen JH. Can Exercise Positively Influence the Intervertebral Disc? Sports Med. Apr 2016;46(4):473–85. doi:10.1007/s40279-015-0444-2

15. Wilke HJ, Neef P, Caimi M, Hoogland T, Claes LE. New in vivo measurements of pressures in the intervertebral disc in daily life. Spine (Phila Pa 1976). Apr 15 1999;24(8):755–62. doi:10.1097/00007632-199904150-00005

16. Min SK, Nakazato K, Yamamoto Y, et al. Cartilage intermediate layer protein gene is associated with lumbar disc degeneration in male, but not female, collegiate athletes. Am J Sports Med. Dec 2010;38(12):2552–7. doi:10.1177/0363546510376714

17. Rottier TD, Allen SJ. The influence of swing leg technique on maximum running speed. J Biomech. Sep 20 2021;126:110640. doi:10.1016/j.jbiomech.2021.110640

18. Kuroki T. The prevention effect of therapeutic exercise on low back pain in athletes. J Phys Med. 1996;7:92.

19. Pfirrmann CW, Metzdorf A, Zanetti M, Hodler J, Boos N. Magnetic resonance classification of lumbar intervertebral disc degeneration. Spine (Phila Pa 1976). Sep 1 2001;26(17):1873–8. doi:10.1097/00007632-200109010-00011

20. Kubo Y, Koyama K, Kimura T. Relationship Between the Anteroposterior Acceleration of Lower Lumbar Spine and Pelvic Tilt Movements During Running. Biomechanics. 2024;4(4):765–772.

21. Tyrrell A, Reilly T, Troup J. Circadian variation in stature and the effects of spinal loading. Spine. 1985;10(2):161–164.

22. Belavy DL, Brisby H, Douglas B, et al. Characterization of Intervertebral Disc Changes in Asymptomatic Individuals with Distinct Physical Activity Histories Using Three Different Quantitative MRI Techniques. J Clin Med. Jun 12 2020;9(6)doi:10.3390/jcm9061841

23. Faul F, Erdfelder E, Buchner A, Lang AG. Statistical power analyses using G*Power 3.1: tests for correlation and regression analyses. Behav Res Methods. Nov 2009;41(4):1149–60. doi:10.3758/BRM.41.4.1149

24. Wilke H, Neef P, Hinz B, Seidel H, Claes L. Intradiscal pressure together with anthropometric data--a data set for the validation of models. Clin Biomech (Bristol, Avon). 2001;16 Suppl 1:S111–26. doi:10.1016/s0268-0033(00)00103-0

25. Meadows KD, Peloquin JM, Newman HR, Cauchy PJK, Vresilovic EJ, Elliott DM. MRI-based measurement of in vivo disc mechanics in a young population due to flexion, extension, and diurnal loading. JOR Spine. Mar 2023;6(1):e1243. doi:10.1002/jsp2.1243

26. Kim J, Yang SJ, Kim H, et al. Effect of shear force on intervertebral disc (IVD) degeneration: an in vivo rat study. Ann Biomed Eng. Sep 2012;40(9):1996–2004. doi:10.1007/s10439-012-0570-z

27. Xia DD, Lin SL, Wang XY, et al. Effects of shear force on intervertebral disc: an in vivo rabbit study. Eur Spine J. Aug 2015;24(8):1711–9. doi:10.1007/s00586-015-3816-2

28. Koyama K, Nakazato K, Min SK, et al. Anterior Limbus Vertebra and Intervertebral Disk Degeneration in Japanese Collegiate Gymnasts. Orthop J Sports Med. Aug 2013;1(3):2325967113500222. doi:10.1177/2325967113500222

29. Sward L, Hellstrom M, Jacobsson B, Nyman R, Peterson L. Disc degeneration and associated abnormalities of the spine in elite gymnasts. A magnetic resonance imaging study. Spine (Phila Pa 1976). Apr 1991;16(4):437–43. doi:10.1097/00007632-199104000-00009

30. Vanhelst J, Zunquin G, Theunynck D, Mikulovic J, Bui-Xuan G, Beghin L. Equivalence of accelerometer data for walking and running: treadmill versus on land. J Sports Sci. May 2009;27(7):669–75. doi:10.1080/02640410802680580

31. Perpiñá-Martínez S, Arguisuelas-Martínez MD, Pérez-Domínguez B, Nacher-Moltó I, Martínez-Gramage J. Differences between Sexes and Speed Levels in Pelvic 3D Kinematic Patterns during Running Using an Inertial Measurement Unit (IMU). International Journal of Environmental Research and Public Health. 2023;20(4):3631.

