## Supplementary Table1 for "Running acceleration correlates with T2 magnetic resonance imaging values of the lumber intervertebral disc"

**Supplementary Table1. Intrarater Reliability of T2 relaxation times analysis**

| ICC(1,1) | ROI1 | ROI2 | ROI3 | ROI4 | ROI5 |
| --- | --- | --- | --- | --- | --- |
| L3-4 |  |  |  |  |  |
| pre | 0.90 | 0.96 | 0.98 | 0.95 | 0.88 |
| post | 0.95 | 0.84 | 0.89 | 0.89 | 0.91 |
| post30 | 0.96 | 0.94 | 0.97 | 0.92 | 0.89 |
| L4-5 |  |  |  |  |  |
| pre | 0.96 | 0.98 | 0.99 | 0.97 | 0.89 |
| post | 0.97 | 0.98 | 1.00 | 0.98 | 0.96 |
| post30 | 0.97 | 0.93 | 0.99 | 0.95 | 0.96 |
| L5-S1 |  |  |  |  |  |
| pre | 0.98 | 0.98 | 0.99 | 0.97 | 0.91 |
| post | 0.95 | 0.98 | 0.99 | 0.95 | 0.91 |
| post30 | 0.98 | 0.84 | 1.00 | 0.98 | 0.91 |

ICC, intraclass correlation coefficient.

The test-retest reliability is shown in Table. The intraclass correlation coefficient values ranged from 0.84 to 1.00, suggesting that the measurement methods used in this study are highly reproducible.
